# Incidentally discovered Covert Cerebrovascular Disease by CT versus MRI: Agreement and Prognostic Value for Stroke and Dementia in a Large Real-World Cohort

**DOI:** 10.1101/2025.09.05.25335192

**Authors:** David M. Kent, Eric J. Puttock, Lester Y Leung, Navdeep Sangha, Neel Madan, Eric E Smith, Mai N Nguyen-Huynh, Wansu Chen

## Abstract

**Background:** Covert cerebrovascular disease (CCD), comprising covert brain infarction (CBI) and white matter disease (WMD), is common in older adults and linked to increased risk of stroke and dementia. While most CCD research relies on MRI, CT remains the predominant imaging modality in clinical care. The influence of imaging modality on detection and prognosis of incidentally discovered CCD remains unclear.

**Methods:** We identified 18,626 patients aged ≥50 years from Kaiser Permanente Southern California who underwent both CT and MRI brain scans within 30 days between 2009– 2022. Patients with known prior stroke or dementia were excluded. Natural language processing algorithms were applied to radiology reports to identify CBI and WMD status and WMD severity (none, mild, moderate, severe). We assessed prevalence, cross-modality agreement (Cohen’s kappa), and reclassification patterns. Prognostic associations with incident stroke or dementia were estimated using Cox Proportional Hazards regression adjusted for vascular and cognitive risk factors.

**Findings:** CBI prevalence was similar for CT (6.3%) and MRI (6.1%), but agreement was modest (κ=0.27). WMD was reported far more often on MRI (60.5%) than CT (24.4%). Among 15,551 patients with classifiable severity on both modalities, 47.9% (n=7,441) had discordant grades, with 92.3% upgraded on MRI. The incidence rates of stroke or dementia per 1,000 person-years were 12.7 (95% CI 11.5 - 14.0) for patients without WMD on either modality (36.3% of the cohort), 22.6 (21.0 - 24.2) for WMD detected on MRI only (39.2% of the cohort), and 52.2 (48.69 to 55.95) for WMD detected on both CT and MRI (21.2% of the cohort). In adjusted Cox models, WMD detected on MRI only was associated with a 23% higher hazard of stroke or dementia (HR=1.23, 95% CI 1.07–1.41) compared with no WMD on either modality, while WMD detected on both CT and MRI was associated with an 82% higher hazard (1.82, 1.58–2.11).

**Interpretation:** MRI detects substantially more WMD than CT; however, WMD visible on CT has stronger prognostic significance, despite CT’s low sensitivity. These findings emphasize modality-based diagnostic and prognostic differences and support the need for modality-specific approaches when translating CCD research into clinical risk assessment and patient counselling.

**Funding:** This work is funded by an Alzheimer’s Drug Discovery Foundation (ADDF) award (RC-202209-2024187) and National Institutes of Health (NIH) grants (2 RF1 NS102233-05, R01 NS134859-01).

**Research in context:** *Evidence before this study:* Most knowledge about covert cerebrovascular disease (CCD), including covert brain infarction (CBI) and white matter disease (WMD), comes from MRI-based research cohorts. However, CT is the dominant imaging modality in routine care, where incidental CCD is identified. Previous studies identified from our literature search suggest that CT and MRI may differ in sensitivity, but no studies have directly compared diagnostic and prognostic differences across both modalities within the same patients in routinely-obtained neuroimaging.

*Added value of this study:* This is the first large-scale, within-subject analysis of CT and MRI CCD findings obtained in routine care. We found that MRI detects substantially more WMD, while CT-detected WMD is more strongly associated with future stroke and dementia. CBI prevalence was similar across modalities, but agreement was modest.

*Implications of all the available evidence:* Our findings suggest that clinically meaningful CCD is usually captured by CT scan; pragmatic research studies need to incorporate this modality.

## Introduction

Covert cerebrovascular disease (CCD), comprising both covert brain infarction (CBI) and white matter disease (WMD), is highly prevalent in older adults. In population-based research cohorts, both CBI and WMD have been consistently associated with elevated risk of future neurological morbidity, including stroke, cognitive decline and dementia.^1-5^ Despite the frequency and clinical impact of these conditions, there are currently no guideline-based treatments for incidentally detected CCD in routine clinical care.

A major obstacle in translating epidemiologic findings into clinical practice is the difference in how CCD is detected in research versus clinical settings. Large-scale research studies typically rely on protocol-driven, screening brain magnetic resonance imaging (MRI) to identify and characterize CCD. In contrast, clinical neuroimaging is generally prompted by acute symptoms (e.g., headaches, dizziness, trauma), with computed tomography (CT) as the first-line modality. This mismatch in patient selection and modality creates a translational gap: CCD patterns well characterized in MRI-based research are often detected under very different circumstances and with different modalities in real-world care.

Variation in detection methods may carry important prognostic consequences. Recent work using natural language processing (NLP) to analyze radiology reports from large healthcare systems suggests that incidentally discovered CCD may carry different prognostic significance when discovered by CT versus MRI.^6-8^ For example, in these studies, even mild WMD detected by CT was associated with risks of future stroke or dementia comparable to, or greater than, those associated with severe WMD on MRI, suggesting that CT may preferentially detect more advanced disease. In contrast, mild WMD on MRI has been associated with outcomes similar to those seen in patients with normal CT scans. However, patients imaged by CT and MRI are quite different in their clinical characteristics, which may also contribute to the observed differences in prognosis. Despite the clinical and research significance of these issues, to our knowledge, no studies have investigated within-subject, cross-modality agreement using clinically indicated, routinely obtained neuroimaging.

To address this gap, we applied a validated NLP algorithm^9,10^ to a cohort of patients who underwent both CT and MRI within a 30-day period. Our objectives were to: (1) compare the prevalence of CBI, WMD, and WMD severity (mild, moderate, severe) across modalities; (2) quantify cross-modality agreement and diagnostic reclassification; and (3) assess the prognostic implications of concordant and discordant findings for future stroke and dementia risk.

## Methods

This study was conducted within the Kaiser Permanente Southern California (KPSC) health system, a large, integrated healthcare organization providing care to over 4.9 million members across 16 hospitals and more than 240 medical offices. The study was approved by the Tufts and KPSC Institutional Review Boards, with a waiver of informed consent.

### Study Population

We included adults aged 50 years or older who were enrolled in the KPSC health plan and underwent both a CT and MRI brain scan within a 30-day interval between 2009 and 2022 (Figure 1). The order of imaging did not affect eligibility; the first scan served as the index modality.

**Figure 1:**
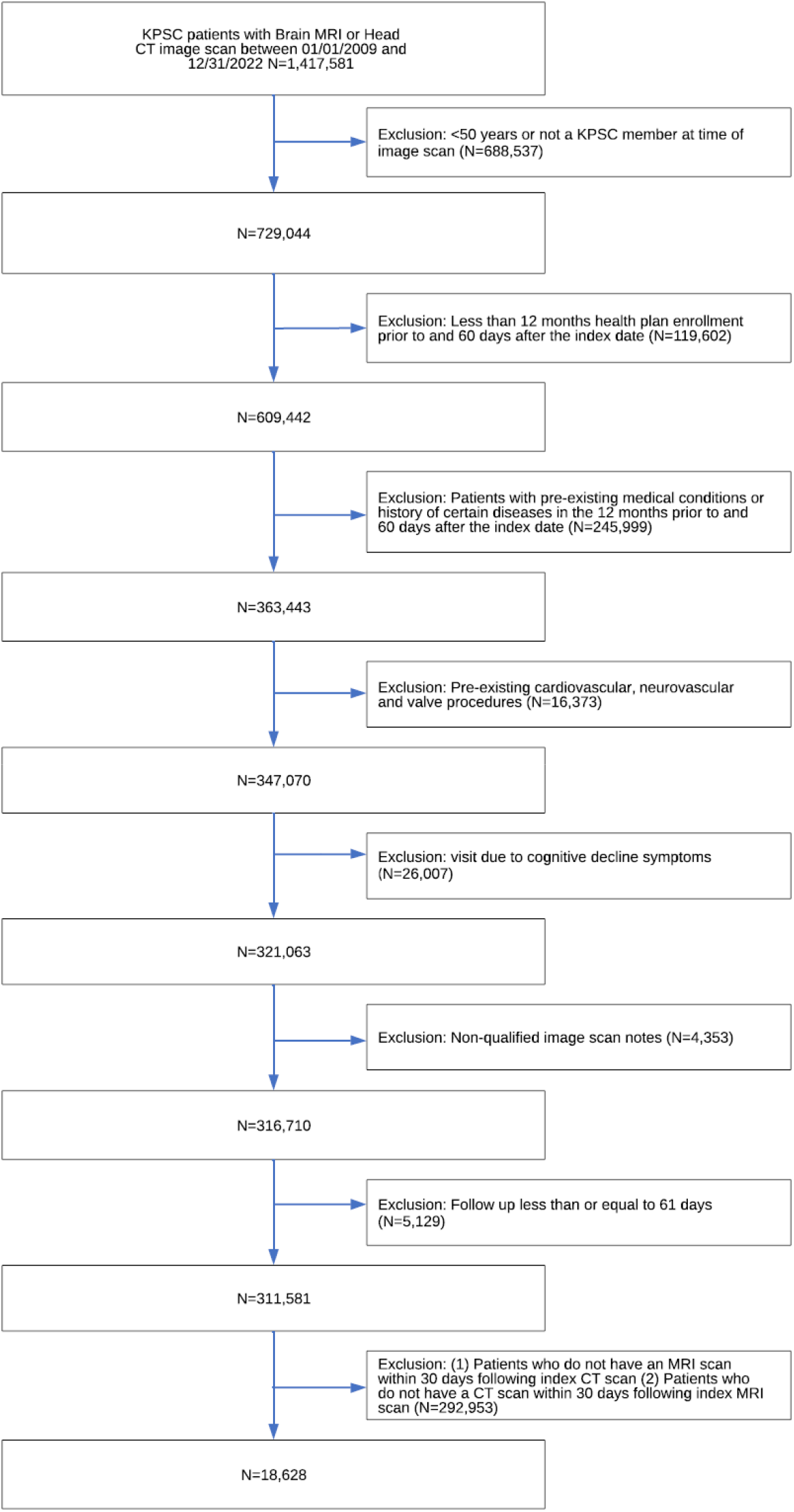
Consort diagram.

To restrict the study population to patients with covert CBI and WMD, we excluded patients with a documented history of ischemic stroke, dementia, or Alzheimer’s disease prior to the index scan. We further excluded individuals whose imaging was obtained for evaluation of acute neurologic symptoms suggestive of major stroke (e.g., transient ischemic attack, hemiplegia, or hemiparesis). The complete ICD-9, ICD-10, and Current Procedural Terminology codes used for exclusions are provided in Technical Appendix Table 1. ^7^ For the present analysis, we also excluded patients with a visit reason or scan indication suggestive of cognitive symptoms or decline (e.g., confusion, disorientation, altered mental status, or dementia evaluation) (Tables 1A and 1B).

**Table 1:**
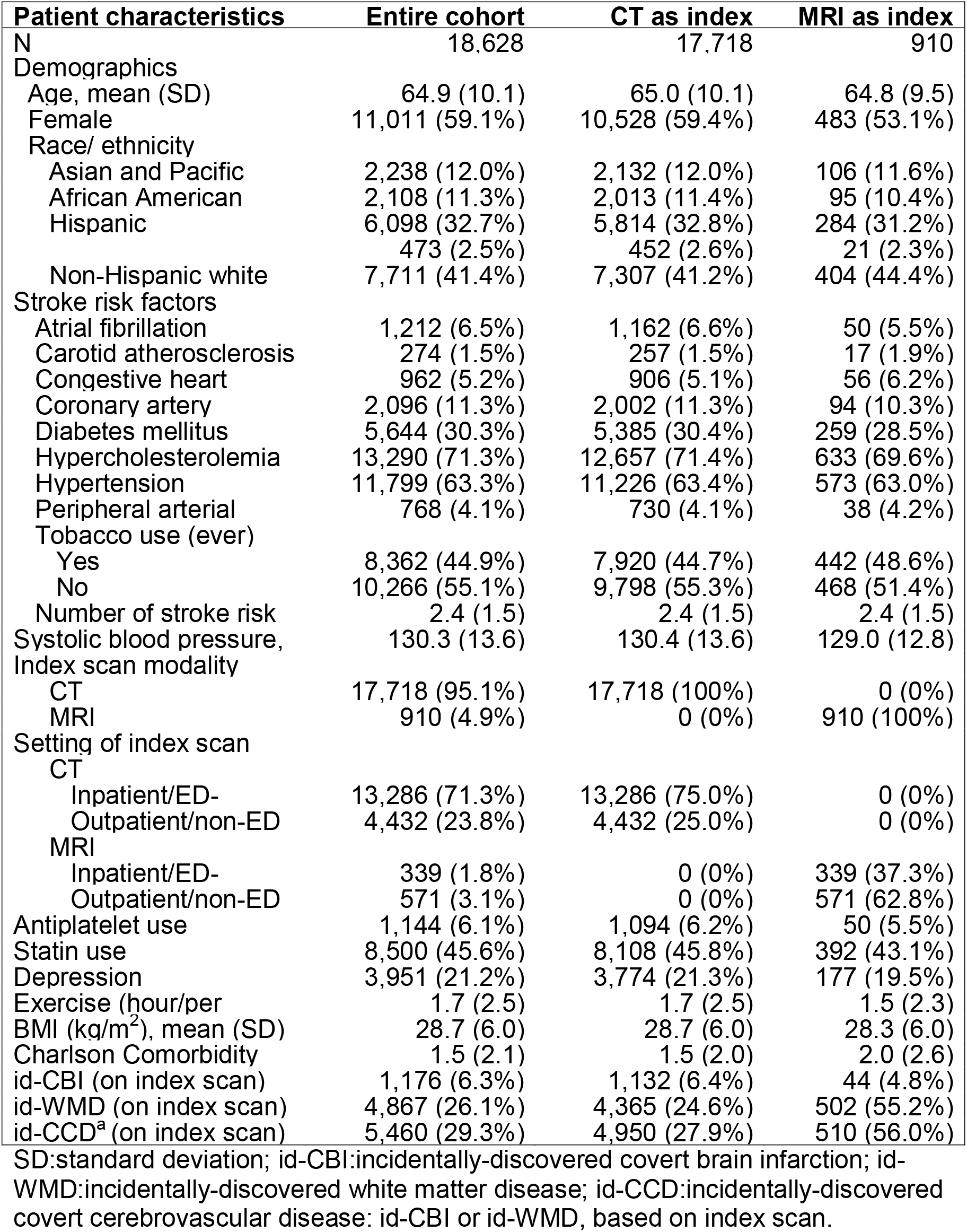
Patient demographic and clinical characteristics at baseline.

| **Patient characteristics** | **Entire cohort** | **CT as index** | **MRI as index** |
| --- | --- | --- | --- |
| N | 18,628 | 17,718 | 910 |
| Demographics |  |  |  |
| Age, mean (SD) | 64.9 (10.1) | 65.0 (10.1) | 64.8 (9.5) |
| Female | 11,011 (59.1%) | 10,528 (59.4%) | 483 (53.1%) |
| Race/ ethnicity |  |  |  |
| Asian and Pacific | 2,238 (12.0%) | 2,132 (12.0%) | 106 (11.6%) |
| African American | 2,108 (11.3%) | 2,013 (11.4%) | 95 (10.4%) |
| Hispanic | 6,098 (32.7%) | 5,814 (32.8%) | 284 (31.2%) |
|  | 473 (2.5%) | 452 (2.6%) | 21 (2.3%) |
| Non-Hispanic white | 7,711 (41.4%) | 7,307 (41.2%) | 404 (44.4%) |
| Stroke risk factors |  |  |  |
| Atrial fibrillation | 1,212 (6.5%) | 1,162 (6.6%) | 50 (5.5%) |
| Carotid atherosclerosis | 274 (1.5%) | 257 (1.5%) | 17 (1.9%) |
| Congestive heart | 962 (5.2%) | 906 (5.1%) | 56 (6.2%) |
| Coronary artery | 2,096 (11.3%) | 2,002 (11.3%) | 94 (10.3%) |
| Diabetes mellitus | 5,644 (30.3%) | 5,385 (30.4%) | 259 (28.5%) |
| Hypercholesterolemia | 13,290 (71.3%) | 12,657 (71.4%) | 633 (69.6%) |
| Hypertension | 11,799 (63.3%) | 11,226 (63.4%) | 573 (63.0%) |
| Peripheral arterial | 768 (4.1%) | 730 (4.1%) | 38 (4.2%) |
| Tobacco use (ever) |  |  |  |
| Yes | 8,362 (44.9%) | 7,920 (44.7%) | 442 (48.6%) |
| No | 10,266 (55.1%) | 9,798 (55.3%) | 468 (51.4%) |
| Number of stroke risk | 2.4 (1.5) | 2.4 (1.5) | 2.4 (1.5) |
| Systolic blood pressure, | 130.3 (13.6) | 130.4 (13.6) | 129.0 (12.8) |
| Index scan modality |  |  |  |
| CT | 17,718 (95.1%) | 17,718 (100%) | 0 (0%) |
| MRI | 910 (4.9%) | 0 (0%) | 910 (100%) |
| Setting of index scan |  |  |  |
| CT |  |  |  |
| Inpatient/ED- | 13,286 (71.3%) | 13,286 (75.0%) | 0 (0%) |
| Outpatient/non-ED | 4,432 (23.8%) | 4,432 (25.0%) | 0 (0%) |
| MRI |  |  |  |
| Inpatient/ED- | 339 (1.8%) | 0 (0%) | 339 (37.3%) |
| Outpatient/non-ED | 571 (3.1%) | 0 (0%) | 571 (62.8%) |
| Antiplatelet use | 1,144 (6.1%) | 1,094 (6.2%) | 50 (5.5%) |
| Statin use | 8,500 (45.6%) | 8,108 (45.8%) | 392 (43.1%) |
| Depression | 3,951 (21.2%) | 3,774 (21.3%) | 177 (19.5%) |
| Exercise (hour/per | 1.7 (2.5) | 1.7 (2.5) | 1.5 (2.3) |
| BMI (kg/m<sup>2</sup>), mean (SD) | 28.7 (6.0) | 28.7 (6.0) | 28.3 (6.0) |
| Charlson Comorbidity | 1.5 (2.1) | 1.5 (2.0) | 2.0 (2.6) |
| id-CBI (on index scan) | 1,176 (6.3%) | 1,132 (6.4%) | 44 (4.8%) |
| id-WMD (on index scan) | 4,867 (26.1%) | 4,365 (24.6%) | 502 (55.2%) |
| id-CCD<sup>a</sup> (on index scan) | 5,460 (29.3%) | 4,950 (27.9%) | 510 (56.0%) |

SD:standard deviation; id-CBI:incidentally-discovered covert brain infarction; id-WMD:incidentally-discovered white matter disease; id-CCD:incidentally-discovered covert cerebrovascular disease: id-CBI or id-WMD, based on index scan.

### Imaging Modality and Exposure Definition

CBI refers to a brain lesion, detected on neuroimaging (MRI or CT), that is believed to result from vascular occlusion in individuals who have no clinical evidence of stroke. WMD is characterized by reduced white matter density on brain CT, or by hyperintense signals on T2-weighted, proton-density, and fluid-attenuated inversion recovery (FLAIR) MRI sequences.

For each modality, the presence of CBI and WMD, and the severity of WMD severity, were classified using a natural language processing (NLP) algorithm applied to radiology reports. The NLP algorithm was applied separately to CT and MRI reports such that the CCD classifications were modality specific.

For outcome analyses, CCD exposure was defined from the first (index) scan: the NLP-derived CBI and WMD classifications on the index scan served as the primary exposure. In secondary analyses we used a modality-specific composite based on both scans (neither, MRI-only, CT-only, both).

#### NLP-Based Classification of WMD and CBI

We applied the NLP algorithm developed at Tufts Medical Center and Mayo Clinic.^9,10^ CBI was classified as positive by NLP if the report language indicated high levels of certainty (e.g., “there is a chronic infarct in the left thalamus” or “likely secondary to a subacute infarct”), but negative if phrased as “possible” (e.g., “the lesion could represent a chronic infarct or dilated perivascular space”).

WMD was identified by NLP using a broad set of terms (e.g., “microangiopathic,” “microvascular ischemic disease,” and “white matter hypodensity”). Severity was classified, using an ordinal scale of mild, moderate, and severe, based on descriptive quantification in the report (e.g., “mild” such as “few scattered foci of T2/FLAIR hyperintensities”; “moderate” such as “multiple foci”; and “severe” such as “diffuse, extensive T2/FLAIR hyperintensities”). Topographic descriptors (e.g., “punctate”, “confluent”) did not affect grading.

The NLP algorithm was validated against both expert reading of the neuroimaging report^10^ and direct reads of a sample of images.^9^ For CBI, the NLP-algorithm had an accuracy, sensitivity, specificity, positive predictive value (PPV), and negative predictive value (NPV) of 0.991, 0.925, 1.000, 1.000, and 0.990, respectively, when compared against expert reading of reports. For WMD, these were: 0.928, 0.924, 0.909, 0.933, and 0.921, respectively. The NLP algorithm demonstrated substantial agreement with direct neuroradiologist image review for WMD severity (κ = 0.52 for WMD severity; F1-score 0.841).

### Outcome Definitions

The primary outcome was a composite of ischemic stroke or dementia. For stroke, we used a highly specific, previously validated algorithm that includes the following ICD-9 codes: 433.x1, 434.x1, 436.x, 437.1x, 437.9x, 438.x. The PPV of this definition has been reported at 97%.^11,12^ This code specifies TIA with a PPV of 70% or higher.^13^ Equivalent ICD-10 codes used are available in Technical Appendix Table 2.^8^ For dementia, we used a broad definition including Alzheimer’s disease, vascular dementia, and mixed dementia^14,15^ (*ICD 9* codes 290.x, 294.20, or 331.0 and corresponding *ICD*-*10* codes). Only a single code was required if accompanied by a cognitive enhancement medication dispensation, or any 2 codes on different dates (with the second as the event date) if cognitive enhancement medication was not dispensed. Positive predictive value has been reported to be >95% for this definition.^16,17^ Equivalent *ICD*-*9* and *ICD*-*10* codes used and qualifying cognitive enhancement medications are shown in Technical Appendix Table 3.^7^

**Table 2:** Comparison of White Matter Disease Severity on CT vs. MRI in Patients Undergoing Both Modalities Within 30 days.

|  |  | **WMD Severity on MRI** |  |  |  |  |  |
| --- | --- | --- | --- | --- | --- | --- | --- |
| **WMD Severity on CT** | **Value** | **No** | **Mild** | **Moderate** | **Severe** | **Non-specified** | **Total** |
| **No** | Frequency | 6770 | 5040 | 879 | 205 | 1184 | 14078 |
|  | Percent | 36.3 | 27.1 | 4.7 | 1.1 | 6.4 | 75.6 |
|  | Row % | 48.1 | 35.8 | 6.2 | 1.46 | 8.4 |  |
|  | Col % | 92.0 | 74.9 | 46.8 | 31.3 | 59.3 |  |
| **Mild** | Frequency | 341 | 1088 | 510 | 147 | 360 | 2446 |
|  | Percent | 1.8 | 5.8 | 2.7 | 0.8 | 1.9 | 13.1 |
|  | Row % | 13.9 | 44.5 | 20.9 | 6.0 | 14.7 |  |
|  | Col % | 4.6 | 16.2 | 27.1 | 22.4 | 18.0 |  |
| **Moderate** | Frequency | 32 | 95 | 169 | 84 | 60 | 440 |
|  | Percent | 0.2 | 0.5 | 0.9 | 0.5 | 0.3 | 2.4 |
|  | Row % | 7.3 | 21.6 | 38.4 | 19.1 | 13.6 |  |
|  | Col % | 0.4 | 1.4 | 9.0 | 12.8 | 3 |  |
| **Severe** | Frequency | 19 | 43 | 46 | 83 | 45 | 236 |
|  | Percent | 0.1 | 0.2 | 0.3 | 0.45 | 0.2 | 1.3 |
|  | Row % | 8.1 | 18.2 | 19.5 | 35.2 | 19.1 |  |
|  | Col % | 0.3 | 0.6 | 2.5 | 12.7 | 2.3 |  |
| **Non-specified** | Frequency | 200 | 467 | 275 | 137 | 349 | 1428 |
|  | Percent | 1.1 | 2.5 | 1.5 | 0.7 | 1.9 | 7.7 |
|  | Row % | 14.0 | 32.7 | 19.3 | 9.6 | 24.4 |  |
|  | Col % | 2.7 | 6.9 | 14.6 | 20.9 | 17.5 |  |
| **Total** | Frequency | 7362 | 6733 | 1879 | 656 | 1998 | 18628 |
|  | Percent | 39.5 | 36.1 | 10.1 | 3.5 | 10.7 | 100 |

**Table 3:** Incident Rates (per 1,000 Person-Years Follow-up) for the Composite Outcome Major Adverse Neurological Events (Stroke or Dementia) and Hazard Ratios Associated with Findings of CBI and WMD on CT and/or MRI

|  | **N** | **Percent** | **Incidence of stroke and dementia** | **Crude hazard ratio** | **Adjusted hazard ratio** |
| --- | --- | --- | --- | --- | --- |
| CBI: neither | 16694 | 89.6 | 21.8 [20.74 to 22.87] | 1 | 1 |
| CBI: MRI only | 754 | 4.2 | 54.8 [47.19 to 63.26] | 2.52 [2.16 to 2.94] | 1.47 [1.23 to 1.76] |
| CBI: CT only | 804 | 4.3 | 43.7 [37.18 to 51.04] | 2.00 [1.69 to 2.36] | 1.26 [1.04 to 1.53] |
| CBI: both | 376 | 2.0 | 64.3 [52.12 to 78.61] | 2.99 [2.42 to 3.70] | 1.68 [1.33 to 2.11] |
| WMD: neither | 6770 | 36.3 | 12.7 [11.50 to 14.01] | 1 | 1 |
| WMD: MRI only | 7308 | 39.2 | 22.6 [21.00 to 24.22] | 1.78 [1.58 to 2.01] | 1.23 [1.07 to 1.41] |
| WMD: CT only | 592 | 3.2 | 37.0 [29.78 to 45.56] | 2.95 [2.34 to 3.73] | 1.46 [1.11 to 1.92] |
| WMD: both | 3958 | 21.3 | 52.2 [48.69 to 55.95] | 4.18 [3.70 to 4.72] | 1.82 [1.58 to 2.11] |

### Statistical Analysis

We calculated the prevalence of CBI and WMD, and the severity distribution of WMD by modality. Agreement between CT and MRI was evaluated using weighted kappa statistics and cross-tabulations. For WMD severity, we assessed the direction of reclassification (none → mild → moderate → severe) among discordant cases and visualized these patterns using river plots. Diagnostic divergence was quantified as the proportion of cases positive on one modality but negative on the other.

Kaplan-Meier curves were plotted to examine stroke and dementia-free survival probability in patients with and without CCD. For WMD, we considered 4 categories: negative on both modalities; MRI-positive/CT-negative; MRI-negative/CT-positive; and positive on both modalities. An analogous classification was used for CBI.

Incidence rates (per 1,000 person-years) and the 95% CIs were calculated using Poisson regression. Cox proportional hazards regression models were used to estimate crude and adjusted hazard ratios (HRs) for the association between modality-specific CCD and outcomes. Adjusted models included established stroke risk factors (age, sex, race/ethnicity, diabetes, hypercholesterolemia, smoking, mean systolic blood pressure [excluding extreme values <70 or >200 mmHg], atrial fibrillation, carotid disease, congestive heart failure, peripheral arterial disease, antiplatelet use, and statin use)^18,19^ and dementia risk factors (depression, body mass index, and exercise).^20-23^

All analyses were conducted using SAS version 9.4 (SAS Institute, Cary, NC).

#### Sensitivity and Stability Analyses

We examined the stability of agreement and diagnostic reclassification to ordering effects by repeating this analysis separately for patients in whom the index scan was a CT and for patients in whom the index scan was an MRI. We also examined the stability of association with outcomes separately across each of the two component outcomes (stroke and dementia) of the primary composite outcome.

## Results

Table 1. presents the demographic and clinical characteristics of the 18,628 patients included in the research cohort. The mean age was 64.9 years (SD 10.1); 59.1% were female. The cohort was racially and ethnically diverse. Cardiovascular risk factors were common: 63.3% had hypertension, 71.3% had hypercholesterolemia, and 30.3% had diabetes; 45.6% were on statin therapy at the time of imaging or in the prior 12 months.

Regarding imaging modality, 95.1% of patients had CT as their index scan, while 4.9% had MRI. The mean time interval between scans was 6.3 days (SD 8.8). The clinical setting differed markedly by modality (**Table 1**): among patients with CT as the index scan, 75.0% underwent imaging in the emergency department or inpatient setting, compared with 37.3% of those with MRI as the index scan. The later were more often imaged in outpatient contexts. **Supplemental Tables 1A and 1B** detail the clinical indications for neuroimaging and the associated reason for visit, respectively.

### Modality-specific prevalence and cross-modality agreement of CCD

Ordering effects did not appear to influence the degree of agreement between CT scan and MRI for either CBI or WMD detection (**Supplemental Tables 2A, 2B, 2C and 2D**) and so results are therefore pooled without regard to the index scan modality.

The prevalence of CBI was similar between CT (6.3%) and MRI (6.1%). Overall agreement on CBI presence or absence was 91.6%, with a Cohen’s kappa of 0.27 (95% CI 0.25-0.30), indicating modest concordance beyond chance. Among patients with CBI on MRI, 33.3% also had CBI on CT; among patients with CBI on CT, 31.9% also had CBI on MRI.

WMD showed a much larger prevalence gap across modalities: 24.4% on CT versus 60.5% on MRI. Agreement for WMD presence was 57.6%, with a Cohen’s kappa of 0.23 (95% CI 0.22 – 0.24). Of patients with WMD on MRI, 35.1% were also positive on CT; of those with WMD on CT, 87.0% were also positive on MRI.

### Classification of WMD severity

Given the absence of scan-order effects on severity grading (**Supplemental Tables 3A and 3B**), analyses were conducted on the pooled dataset.

Among the 15,551 patients with WMD severity classified on both CT and MRI (Table 2), nearly half (n = 7,441; 47.9%) were assigned different severity grades across modalities. Most reclassifications (n = 6,865; 92.3% of reclassified cases) represented an upgrade to a higher severity category on MRI, consistent with MRI’s greater sensitivity to white matter changes (Figure 2).

**Figure 2:**
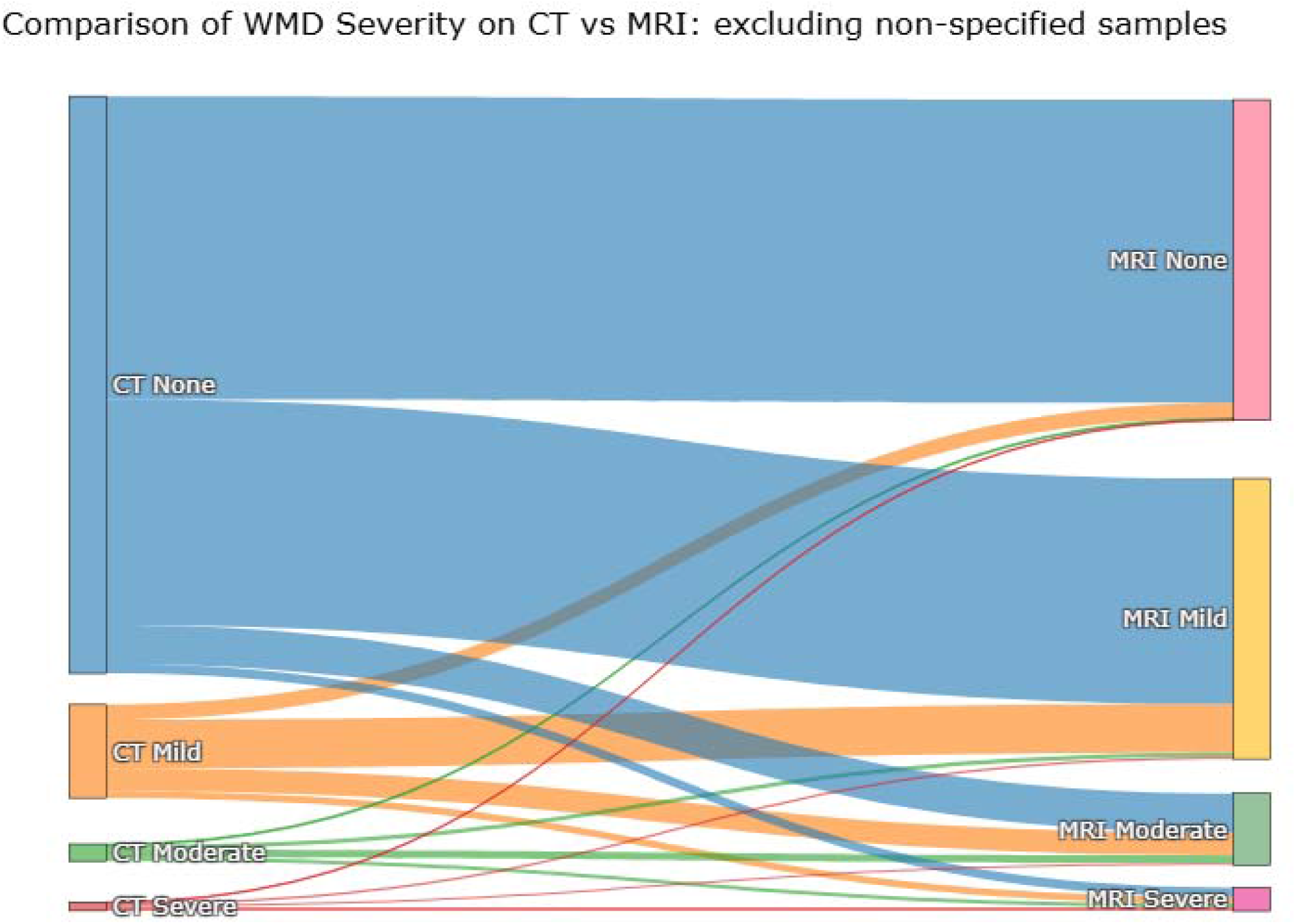
River Plot showing reclassification of white matter disease severity on CT scan and MRI (excluding patients with unspecified severity on either scan) This river plot depicts reclassification of WMD severity on CT scan on the left versus MRI findings on the right. Most patients with incidentally discovered WMD have mild disease, whether identified on CT or on MRI. When reclassification occurs most patients are reclassified to a higher severity class by MRI. The most common type of reclassification were those with no WMD on CT being reclassified as mild WMD on MRI.

Discrepancies were most common when CT reported no WMD. Of the 14,078 CT-negative cases, MRI confirmed no WMD in 6,770 (48.1%), but identified mild WMD (35.8%), moderate (6.2%), or severe (1.5%) WMD in the remainder. Conversely, among the 7,362 MRI-negative cases, CT was also negative in 6,770 (92.0%).

When MRI indicated mild WMD (n = 6,733), CT reported no WMD in 5,040 cases (74.9%), mild WMD in 1,088 (16.2%), and moderate or severe WMD in 138 (2.0%). These cross-classification patterns are shown in Table 2, with corresponding reclassification flows illustrated in Figure 2 (excluding “unspecified severity”) and in Supplementary Figure 1 (including cases with unspecified severity).

Figure 4. shows selected examples of discordant cases, including a patient with findings on MRI but not on CT (Patient A), and one with findings with CT but not on MRI (Patient B).

### Associations between CCD and Stroke and Dementia

The mean follow up was 4.4 years (SD 3.7). During study follow-up, there were 985 persons with stroke events only, 716 persons with dementia events only, and 330 persons with both stroke and dementia events.

For the CBI exposure (Table 3), the incidence rate of stroke or dementia was 21.8 per 1,000 person-years (95% CI, 20.7 - 22.9)) among patients negative on both CT and MRI (89.6% of the cohort) and 64.3 per 1,000 person-years (52.1 to 78.6) among those positive on both modalities (2.0% of the cohort). Patients with discordant findings had intermediate rates: MRI-only detection (4.2% of patients) was associated with 54.8 per 1,000 person-years (47.2 - 63.3), and CT-only detection (4.3% of patients) with 43.7 per 1,000 person-years (37.2 - 51.0). Crude and adjusted hazard ratios followed the same gradient (Table 3), with MRI-only detection showing a nominally higher risk than CT-only detection.

For the WMD exposure (**Table 3**), the incidence rate of stroke or dementia was 12.7 per 1,000 person-years (11.5 to 14.0) for patients negative on both modalities (36.3% of the cohort) and 52.2 per 1,000 person-years (48.7 to 56.0) for those positive on both (21.2%). Rates for the discordant groups fell between these values, though— unlike with CBI— discordant cases fell predominantly in the MRI only class. MRI-only detection (39.2% of patients) had an incidence rate of 22.6 per 1,000 person-years (21.0 to 24.2), whereas the rare cases with CT-only detection (3.2% of patients) showed a substantially higher rate of 37.0 per 1,000 person-years (29.8 to 45.6). Fully adjusted hazard ratios reflected these differences, with the highest risk in those that had WMD on both MRI and CT (HR 1.82, 95% CI, 1.58–2.11), followed by CT-only (1.46, 1.11–1.92) and MRI-only (1.23, 1.07–1.41) groups.

Kaplan Meier curves showing event-free survival for patients both concordant and discordant for the findings of CCD are shown in **Figure 3a** (for CBI) and in **Figure 3b** (for WMD). **Supplemental Table 4A and 4B** show the incidence rates and hazard ratios for each component of the composite outcome, which show consistency of the modality-specific effects for both stroke and dementia.

**Figure 3:**
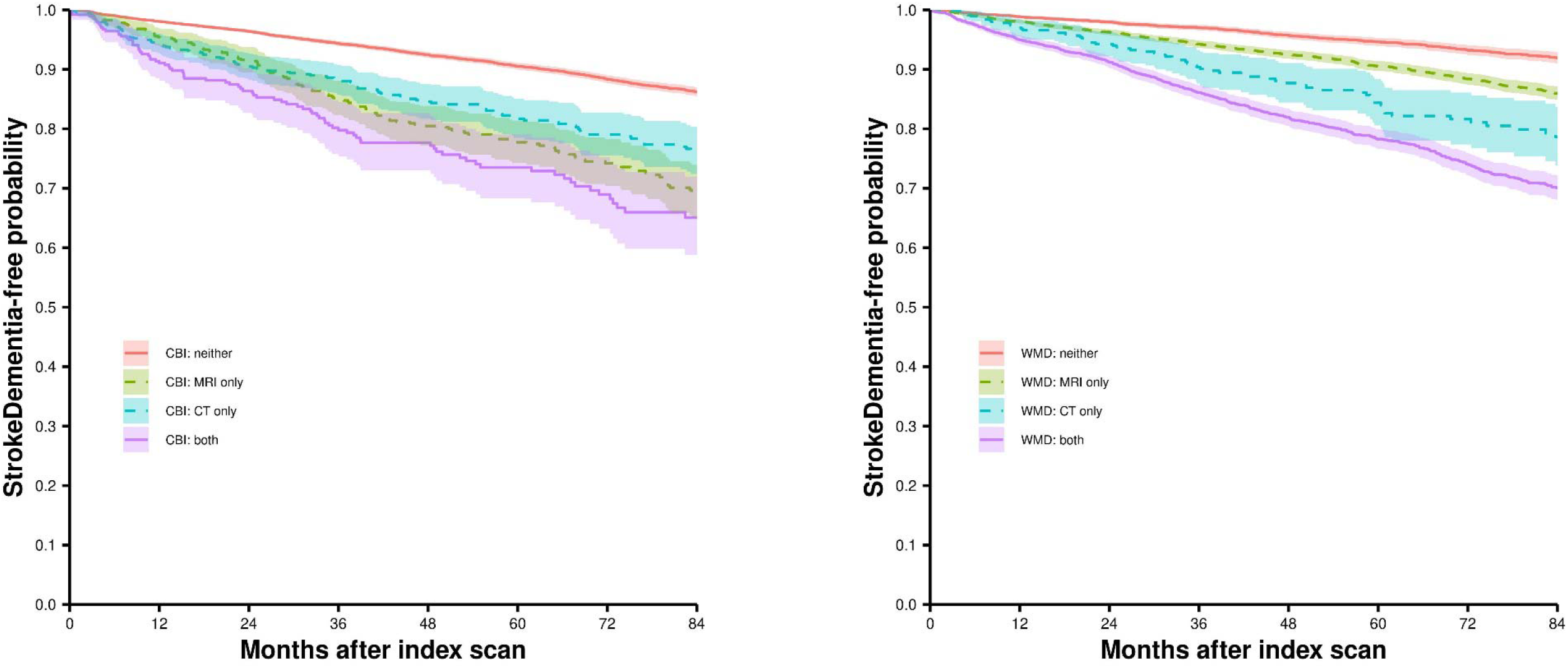
Kaplan Meier Event Free Probability for CBI and WMD exposure groups. This figure depicts the event free survival for patients concordant and discordant for findings of covert cerebrovascular disease on MRI and on CT scan. The plot on the left side shows the event survival for the findings of CBI and the plot on the right shows the event free survival for the findings of WMD. Shadings represent 95% confidence intervals.

**Figure 4:**
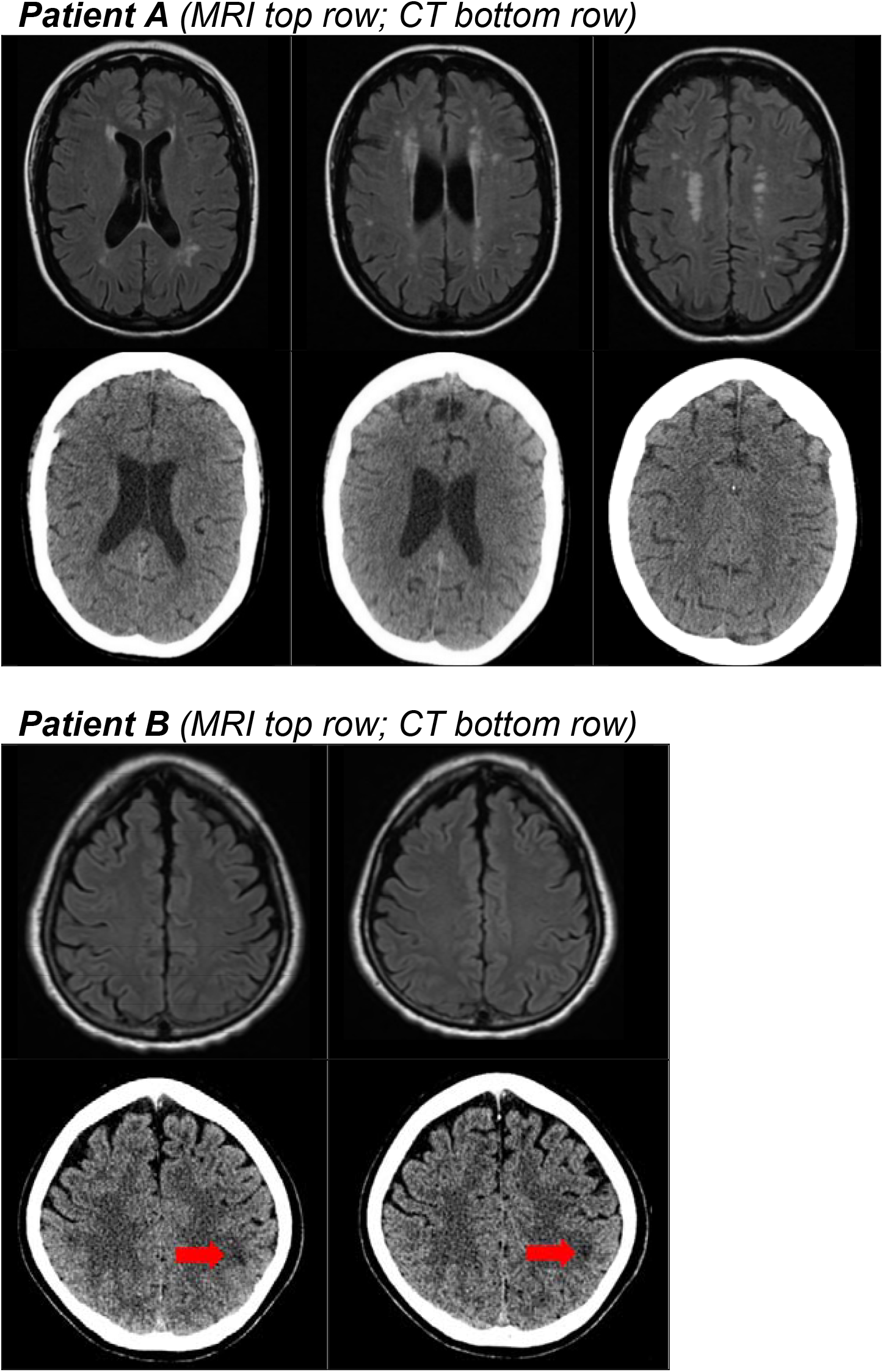
Two examples of discordant MRI and CT scans for the presence of white matter disease. Examples of discordant findings between MRI and CT scans for white matter disease (WMD). For patient A (MRI top row; CT bottom row), WMD is clearly visible on the MRI FLAIR sequence, particularly in the periventricular region, but not on CT. On natural language processing (NLP) interpretation, the MRI was classified as mild WMD, whereas the CT was classified as normal. For patient B (MRI top row; CT bottom row), the MRI FLAIR sequence was interpreted as showing no WMD, while the corresponding CT scan demonstrated juxtacortical white matter changes (red arrow). On NLP interpretation, the MRI was classified as no WMD and the CT as mild WMD.

## Discussion

In this large study of over 18,000 patients over age 50—free of known stroke and dementia at baseline—who underwent both CT and MRI of the brain within a 30-day interval, we found substantial cross-modality discrepancies in CCD detection and WMD severity grading. While CBI prevalence was similar between CT and MRI (approximately 6%), WMD was more than twice as common on MRI. When WMD was classified as mild on MRI, it was absent on the CT report in about three-quarters of cases. Even when MRI reported severe WMD, the corresponding CT report indicated no WMD in nearly one-third of cases. Approximately half of all patients had different WMD severity grades across modalities, with over 90% of those reclassifications assigned a higher severity grade on MRI. This pattern underscores MRI’s ability to detect a broader range of white matter changes, particularly subtle or early-stage lesions that are not apparent on CT.

Prognostically, compared to patients negative for WMD on both modalities, WMD seen on MRI alone was associated with an approximately 20% higher hazard of future stroke or dementia, whereas risk was ∼80% higher when WMD was also detected on CT and MRI. From a clinical standpoint, while no specific evidence-based therapies are available for this condition, our results suggest that CT-identified WMD might prompt optimization of vascular risk factor management. In research, modality-specific case definitions may be warranted when selecting participants for trials targeting small vessel disease to optimize the risk profile of trial enrollees.

MRI, particularly with FLAIR and T2-weighted sequences, is considered the gold standard for detecting cerebral small vessel disease due to its ability to capture subtle, early-stage white matter changes, including clinically silent infarcts and leukoaraiosis.^24- 26^ However, this high sensitivity may also be a disadvantage: MRI detects a broader, more biologically heterogeneous spectrum of lesions, some of which may have little clinical relevance, leading to potential overdiagnosis. Additionally, while MRI might be preferred in research for its assay sensitivity in detecting lesions and subtle changes over time, CT remains far more common in routine care and appears to identify WMD with greater prognostic relevance. In our broader KPSC cohort of 241,050 older, stroke- and dementia-free patients, 74% received CT rather than MRI. Given that roughly 25 brain imaging exams are performed per 1,000 people annually in developed countries, U.S. hospital archives now contain millions of CT scans from older adults. These datasets, when combined with AI-based tools, offer a major opportunity for large-scale, “opportunistic screening” to study CCD and potentially reduce neurological morbidity. In contrast, research based solely on screened MRI cohorts may be less generalizable to these real-world clinical populations, where the great majority of clinically-relevant findings (i.e. those forecasting future stroke and dementia) are detected on CT.

There have been a very limited number of prior studies that have compared MRI to CT scan cross-sectionally in the same individuals for findings suggestive of small vessel disease.^27-29^ These studies are several decades old, using prior generations of technology and were small, limited to about 20 patients. Nevertheless, they consistently conclude MRI to have a much higher sensitivity for these findings. Interestingly, one of these studies, which compared CT and MRI WMD findings in 27 patients from a memory-clinic, found a similarly higher comparative prevalence of WMD in MRI versus CT as the current study, and also found that only CT-detected WMD was significantly associated with the development of abnormal neurological signs (such as abnormal gait or asymmetric reflexes) over one year of follow up.

In some contrast both to our study and these earlier ones, a study of 70 acute stroke patients from the International Stroke Trial-3 using visual rating scales of WMD and atrophy emphasized that MRI and CT findings were fairly well correlated, particularly for periventricular areas, although FLAIR’s higher sensitivity reduced agreement.^30^ Nevertheless, MRI remained more sensitive than CT for WMD and lacunar infarcts. The somewhat higher kappa values reported in this study likely reflect a highly controlled research setting, with only two readers with extensive pre-study training and coaching, an approach that may not scale to either real world practice or large-scale research.

While aligned with these prior studies, our study is unique in many respects: it leveraged a very large diverse population of patients receiving clinically indicated scans and was several orders of magnitude larger than other studies; it specifically targeted CCD; it employed a pragmatic approach examining neuroimaging reports reflective of routine care; it is based on contemporaneous imaging technology; it examined disagreement not only for the presence of findings but in the severity of WMD; and it is the only cross-modality study to our knowledge that followed patients over time to examine the associations of these findings with the hard clinical outcomes of stroke and dementia, both in patients with concordant and discordant findings across modalities. Thus, our study led to unique insights on the trade-off between the two main neuroimaging modalities in identifying clinically relevant CCD.

Several limitations warrant consideration. First, our findings here are based on the subset of patients that received both clinically indicated CT and clinically indicated MRI scans. These patients represent a selected subset (less than 10%) of our overall neuroimaging cohort and may not be representative of other patients receiving clinically indicated scans. Second, we relied on NLP review of radiology reports rather than direct image review. Although our algorithm has been validated with high accuracy for CBI, and for WMD presence and severity, some discrepancies may reflect variability in radiologist reporting or NLP misclassification. However, our findings are likely to reflect the associations found in routine care, improving generalizability to routine practice. Additionally, the consistent pattern—greater sensitivity of MRI but stronger prognostic associations for CT-detected disease—is unlikely to be an artifact of reporting or NLP methods. We note the cases with the surprising finding of WMD on CT but not MRI (which comprise only 3% of the total sample) may arise in part from a combination of neuroradiologist overcalling disease on CT and underreporting visible changes on MRI, as well as NLP error. However, on direct review of a sample of scans, we did find scans with clearly visible changes on CT and normal MRIs (**Figure 4**), and the relatively high risk of stroke and dementia in this group suggests that this is clinically-relevant disease.

In summary, MRI identifies WMD more frequently and assigns higher severity than CT, but WMD detected by CT is more strongly linked to future stroke and dementia. These findings have important implications for risk stratification, clinical communication, and the translation of imaging-based research into practice. Future studies should aim to harmonize WMD severity classification across modalities and explore whether modality-specific thresholds can better align prognostic models with real-world imaging practices.

## Data Availability

The datasets generated or analyzed during the current study are not publicly available due to ethical standards. The authors do not have permission to share data.

## Reference list

1. Debette S, Schilling S, Duperron MG, Larsson SC, Markus HS. Clinical Significance of Magnetic Resonance Imaging Markers of Vascular Brain Injury: A Systematic Review and Meta-analysis. JAMA Neurol 2019; 76(1): 81–94.

2. Debette S, Markus HS. The clinical importance of white matter hyperintensities on brain magnetic resonance imaging: systematic review and meta-analysis. BMJ 2010; 341: c3666.

3. Vermeer SE, Hollander M, van Dijk EJ, Hofman A, Koudstaal PJ, Breteler MM. Silent brain infarcts and white matter lesions increase stroke risk in the general population: the Rotterdam Scan Study. Stroke 2003; 34(5): 1126–9.

4. Vermeer SE, Prins ND, den Heijer T, Hofman A, Koudstaal PJ, Breteler MM. Silent brain infarcts and the risk of dementia and cognitive decline. NEJM 2003; 348(13): 1215–22.

5. Sigurdsson S, Aspelund T, Kjartansson O, et al. Incidence of Brain Infarcts, Cognitive Change, and Risk of Dementia in the General Population: The AGES-Reykjavik Study (Age Gene/Environment Susceptibility-Reykjavik Study). Stroke 2017; 48(9): 2353–60.

6. Wang AY, Leung LY, Puttock EJ, et al. Stratifying future stroke risk with incidentally-discovered white matter disease severity and covert brain infarct site. Cerebrovasc Dis 2023.

7. Kent DM, Leung LY, Zhou Y, et al. Association of Incidentally Discovered Covert Cerebrovascular Disease Identified Using Natural Language Processing and Future Dementia. J Am Heart Assoc 2023; 12(1): e027672.

8. Kent DM, Leung LY, Zhou Y, et al. Association of silent cerebrovascular disease identified using natural language processing and future ischemic stroke. Neurology 2021; 97(13): e1313–e21.

9. Leung LY, Fu S, Luetmer PH, et al. Agreement between neuroimages and reports for natural language processing-based detection of silent brain infarcts and white matter disease. BMC Neurol 2021; 21(1): 189.

10. Fu S, Leung LY, Wang Y, et al. Natural Language Processing for the Identification of Silent Brain Infarcts From Neuroimaging Reports. JMIR Med Inform 2019; 7(2): e12109.

11. Birman-Deych E, Radford MJ, Nilasena DS, Gage BF. Use and effectiveness of warfarin in Medicare beneficiaries with atrial fibrillation. Stroke 2006; 37(4): 1070–4.

12. Andrade SE, Harrold LR, Tjia J, et al. A systematic review of validated methods for identifying cerebrovascular accident or transient ischemic attack using administrative data. Pharmacoepidemiology and drug safety 2012; 21 Suppl 1(Suppl 1): 100–28.

13. Kokotailo RA, Hill MD. Coding of stroke and stroke risk factors using international classification of diseases, revisions 9 and 10. Stroke 2005; 36(8): 1776–81.

14. Taylor DH, Jr., Fillenbaum GG, Ezell ME. The accuracy of medicare claims data in identifying Alzheimer’s disease. J Clin Epidemiol 2002; 55(9): 929–37.

15. Taylor DH, Jr., Østbye T, Langa KM, Weir D, Plassman BL. The accuracy of Medicare claims as an epidemiological tool: the case of dementia revisited. J Alzheimers Dis 2009; 17(4): 807–15.

16. Quan H, Li B, Saunders LD, et al. Assessing validity of ICD-9-CM and ICD-10 administrative data in recording clinical conditions in a unique dually coded database. Health Serv Res 2008; 43(4): 1424–41.

17. van de Vorst IE, Vaartjes I, Sinnecker LF, Beks LJ, Bots ML, Koek HL. The validity of national hospital discharge register data on dementia: a comparative analysis using clinical data from a university medical centre. Neth J Med 2015; 73(2): 69–75.

18. Wolf PA, D’Agostino RB, Belanger AJ, Kannel WB. Probability of stroke: a risk profile from the Framingham Study. Stroke 1991; 22(3): 312–8.

19. Flueckiger P, Longstreth W, Herrington D, Yeboah J. Revised Framingham Stroke Risk Score, Nontraditional Risk Markers, and Incident Stroke in a Multiethnic Cohort. Stroke 2018; 49(2): 363–9.

20. Yan S, Fu W, Wang C, et al. Association between sedentary behavior and the risk of dementia: a systematic review and meta-analysis. Transl Psychiatry 2020; 10(1): 112.

21. Chan YE, Chen MH, Tsai SJ, et al. Treatment-Resistant depression enhances risks of dementia and alzheimer’s disease: A nationwide longitudinal study. J Affect Disord 2020; 274: 806–12.

22. Barnes DE, Covinsky KE, Whitmer RA, Kuller LH, Lopez OL, Yaffe K. Dementia risk indices: A framework for identifying individuals with a high dementia risk. Alzheimers Dement 2010; 6(2): 138–41.

23. Qizilbash N, Gregson J, Johnson ME, et al. BMI and risk of dementia in two million people over two decades: a retrospective cohort study. Lancet Diabetes Endocrinol 2015; 3(6): 431–6.

24. Wardlaw JM, Smith EE, Biessels GJ, et al. Neuroimaging standards for research into small vessel disease and its contribution to ageing and neurodegeneration. Lancet Neurol 2013; 12(8): 822–38.

25. Prins ND, Scheltens P. White matter hyperintensities, cognitive impairment and dementia: an update. Nat Rev Neurol 2015; 11(3): 157–65.

26. Smith EE, Barber P, Field TS, et al. Canadian Consensus Conference on Diagnosis and Treatment of Dementia (CCCDTD)5: Guidelines for management of vascular cognitive impairment. Alzheimers Dement (N Y) 2020; 6(1): e12056.

27. Brown JJ, Hesselink JR, Rothrock JF. MR and CT of lacunar infarcts. AJR Am J Roentgenol 1988; 151(2): 367–72.

28. Seiderer M, Krappel W, Moser E, et al. Detection and quantification of chronic cerebrovascular disease: comparison of MR imaging, SPECT, and CT. Radiology 1989; 170(2): 545–8.

29. Lopez OL, Becker JT, Jungreis CA, et al. Computed tomography--but not magnetic resonance imaging--identified periventricular white-matter lesions predict symptomatic cerebrovascular disease in probable Alzheimer’s disease. Arch Neurol 1995; 52(7): 659–64.

30. Ferguson KJ, Cvoro V, MacLullich AMJ, et al. Visual Rating Scales of White Matter Hyperintensities and Atrophy: Comparison of Computed Tomography and Magnetic Resonance Imaging. J Stroke Cerebrovasc Dis 2018; 27(7): 1815–21.

